# Variations in human monocyte-derived macrophage antimicrobial activities and their associations with tuberculosis clinical manifestations

**DOI:** 10.1101/2020.09.14.20194043

**Authors:** Trinh T. B. Tram, Vu T. N. Ha, Do D. A. Thu, Tran D. Dinh, Hoang N. Nhung, Nguyen T. Hanh, Nguyen H. Phu, Guy E. Thwaites, Nguyen T. T. Thuong

**Affiliations:** Oxford University Clinical Research Unit, Ho Chi Minh City, Vietnam; Centre for Tropical Medicine and Global Health, Nuffield Department of Medicine, University of Oxford, Oxford, United Kingdom

**Author notes:** **Correspondence:** Nguyen T. T. Thuong.

**Keywords:** tuberculosis, acidification, proteolysis, human macrophage, variation

## Abstract

Macrophages play a significant role in preventing infection through antimicrobial activities, particularly acidification and proteolysis. *Mycobacterium tuberculosis* infection can lead to diverse outcomes, from latent asymptomatic infection to active disease involving multiple organs. Monocyte-derived macrophage is one of the main cell types accumulating in lungs following *Mtb* infection. The variation of intracellular activities of monocyte-derived macrophages in humans and the influence of these activities on the tuberculosis (TB) spectrum are not well understood. By exploiting ligand-specific bead-based assays, we investigated macrophage antimicrobial activities real-time in healthy volunteers (n=53) with 35 cases of latent TB (LTB), and those with active TB (ATB) and either pulmonary TB (PTB, n=70) or TB meningitis (TBM, n=77). We found wide person-to-person variations in acidification and proteolytic activities in response to both non-immunogenic IgG and pathogenic ligands comprising trehalose 6,6′-dimycolate (TDM) from *Mycobacterium tuberculosis* or β-glucan from *Saccharamyces cerevisiase*. The variation in the macrophage activities remained similar regardless of stimuli; however, IgG induced stronger acidification activity than immunogenic ligands TDM (P=10^−5^, 3×10^−5^ and 0.01 at 30, 60 and 90 min) and β-glucan (P=10^−4^, 3×10^−4^ and 0.04 at 30, 60 and 90 min). Variation in proteolysis activity was slightly higher in LTB than in ATB (CV=40% in LTB vs. 29% in ATB, P=0.03). There was no difference in measured antimicrobial activities in response to TDM and bacterial killing in macrophages from LTB and ATB, or from PTB and TBM. Our results indicate that antimicrobial activities of monocyte-derived macrophages vary among individuals and show immunological dependence, but suggest these activities cannot be solely responsible for the control of bacterial replication or dissemination in TB.

## Introduction

Macrophages play an important role in innate immunity to protect the human body from microbial infection. After recognizing pathogens via surface receptors, macrophages phagocytose bacteria into phagosomes. These phagosomes are key to macrophage antimicrobial activities. The phagosomes undergo a maturation process in which they are acidified by recruiting V-ATPases(Levin, Grinstein, and Canton 2016) then sequentially fuse with multiple intra-vesicles including, lastly, lysosomes to form phago-lysosomes. The environment inside these vesicles is highly acidic, around pH 4.5, and enriched with an assortment of peptidases and hydrolases, which can kill the bacteria and process antigens for presentation to T cells, leading on to secondary adaptive immune responses(Flannagan, Cosío, and Grinstein 2009; Levin, Grinstein, and Canton 2016).

Macrophages are thought to exhibit different levels of antimicrobial activities depending on the nature of stimuli, but the pattern of difference is not consistent between different *in vitro* macrophage models (Kuldeep Sachdeva et al. 2020; Podinovskaia et al. 2013; Yates, Hermetter, and Russell 2005). For example, in mouse bone marrow-derived macrophages (BMDM) phagocytosis of *Mycobacterium tuberculosis* (*Mtb*) results in a higher proteolytic activity than phagocytosis of non-immunogenic IgG, but the induced acidification activity is at a similar level(Podinovskaia et al. 2013). However, human monocyte-derived macrophages (MDM) exhibit reduced acidification and proteolytic activities in response to the bacteria or pathogenic ligands(Yates, Hermetter, and Russell 2005; Podinovskaia et al. 2013). These observations are from studies with only one or two human participants, with no attention to the potential variation of antimicrobial activities in human macrophages from different donors in response to the stimuli.

*Mtb* is an intracellular pathogen for which infection leads to diverse outcomes. One quarter of the human population is considered to have been exposed to *Mtb* infection(Cohen et al. 2019), of which 5-10% develop tuberculosis (TB) disease within 2-5 years of infection. The remaining 90% are widely assumed to have latent TB (LTB) infection that usually lasts throughout the person’s life time without any symptoms(Cadena, Fortune, and Flynn 2017). The primary site of active TB infection is the lungs, causing pulmonary TB (PTB); however, the bacteria can disseminate by the bloodstream resulting in TB in other organs(Polena et al. 2016; Wilkinson et al. 2017). Tuberculosis meningitis (TBM) is the most severe form of extra-pulmonary TB, causing death or neurological disability in around 50% of survivors(Mai et al. 2018; Marais et al. 2011).

Host genetics is associated with susceptibility to different clinical manifestations of TB infection but little is known about the underlying mechanism(Berrington and Hawn 2007). For example, a polymorphism in the receptor gene Toll-Like Receptor 2 is preferentially associated with TBM in comparison to pulmonary TB (Thuong et al. 2007). Because *Mtb* recognition by receptors is upstream of the phagocytosis process, the polymorphisms in these genes may influence the phagocytosis and antimicrobial activities, resulting in different outcomes of *Mtb* infection. A previous study shows a polymorphism in macrophage receptor with collagenous structure (MARCO), a receptor for trehalose 6,6′-dimycolate (TDM) from *Mtb*, is associated with impaired phagocytosis, which could lead to defective antimicrobial activities and thus increases susceptibility to active disease (Thuong et al. 2016). These works suggest that macrophage activities, under influence of host genetics, may contribute significantly to determining whether infection results in latent, localized or disseminated disease.

Diversity in macrophage populations which derive from different ontogenies is found in the lungs, the primary sites of *Mtb* infection (S. Y. S. Tan and Krasnow 2016). The distinct capabilities of these populations in controlling *Mtb* growth have emerged recently in studies using a mouse model(Huang et al. 2018). Resident alveolar macrophages present a permissive environment for *Mtb* growth(Huang et al. 2018) and decrease in numbers during the course of infection(Lee et al. 2020). Whereas, interstitial macrophages, derived from blood monocytes, accumulate in lungs, exhibit a Th1-like cell-type and restrict bacterial growth(Huang et al. 2018). Moreover, depletion of MDM results in an increased bacterial burden in mice lungs, suggesting the MDM population plays a central role in determining infection progress. Previous studies demonstrate the influence of phagosomal activities of MDM on restriction of *Mtb* growth. Increased macrophage phagosome maturation leads to more efficient mycobacterial killing in BMDM and is associated with reduced bacterial burden in mice organs(Hostetter et al. 2002; Welin et al. 2011; Härtlova et al. 2018. In a human macrophage model, loss of acidification or protease activity by inhibitors or cell death results in increased *Mtb* replication(Welin et al. 2011; Mahamed et al. 2017). In spite of the strong evidence *in vitro*, there have been few studies investigating the association of variations in antimicrobial activities of MDM with development and progression of TB disease.

Previous studies have measured macrophage antimicrobial activities, for example acidification, by using fluorescent weak bases which accumulate in acidic compartments(Mwandumba et al. 2004; Hostetter et al. 2002; Welin et al. 2011). However these probes are often qualitative, indicating whether the phagosome is acidified or not, and may not be accurate for quantifying the phagosomal acidity(Canton and Grinstein 2015). In addition, these probes are not phagosome-specific since they can also label other acidic cellular organelles such as lysosomes(Canton and Grinstein 2015). In order to overcome these limitations, we previously adopted the bead model of Russell(Russell et al. 2009) and developed ligand-coated beads to measure macrophage antimicrobial activities(Tram et al. 2019). The beads were labeled with reporters for acidification or proteolytic activity, which allowed these activities to be measured accurately in real-time and specifically in the bead-containing phagosomes. Beads can also be coated with non-immunogenic or immunogenic ligands from the pathogen, allowing us to study variation of macrophage antimicrobial activities in response to different ligands and to dissect the specific pathways associated with such variation. This model of bead-treated macrophages also enables us to handle multiple conditions with many experimental replications at the same time using a microplate reader.

In this study, we assessed the variation in antimicrobial activities, primarily acidification and proteolysis, of MDM from 53 Vietnamese healthy volunteers in response to immunogenic or non-immunogenic ligands, using our ligand-coated bead models. We then examined these activities in macrophages from 182 participants with different TB clinical manifestations, comprising LTB, PTB and TBM. We hypothesized that there would be variations in the antimicrobial activities of MDM among the participants in response to different ligands and that these variations would correlate with the control of bacteria and different clinical phenotypes.

## Materials and methods

### Participants

Healthy adult donors and patients with PTB or TBM, who were all HIV negative, were recruited to this study. Healthy donors were recruited from Vietnamese staff working at Oxford University Clinical Research Unit, Vietnam. These participants were further screened for LTB infection using T-SPOT.TB assays (TSPOT.TB, Oxford Immunotec, UK). Thirty-five donors who had positive results were considered to have LTB. PTB patients were recruited at the start of treatment from District TB Units (DTUs) 4 and 8 in Ho Chi Minh City (HCMC), Vietnam from Jan 2015 to Feb 2016 as described previously(Vijay et al. 2017). PTB patients had no previous history of TB treatment, had sputum positive with acid fast bacilli by Ziehl-Neelsen stain and chest X-rays. TBM patients were recruited from a clinical trial conducted at Hospital of Tropical Diseases, HCMC, from Sep 2014 to Feb 2016(Mai et al. 2018). The patients had clinical meningitis, which was defined as the combination of nuchal rigidity and abnormal cerebrospinal fluid parameters, and/or acid fast bacilli seen in the cerebrospinal fluid by Ziehl-Neelsen stain or culture. Peripheral blood was collected on enrollment. These samples were anonymized by subject codes, without names or identifying information. The collection and processing of samples from LTB, PTB and TBM participants were carried out simultaneously, as much as possible, to minimize artificial technical variations in data for the different groups.

Written informed consent was obtained from each participant or an accompanying relative if the participant was unable to provide consent independently. The study protocol and informed consent form were approved by the institutional review boards of the Hospital for Tropical Diseases in Vietnam and the Oxford Tropical Research Ethics Committee in the United Kingdom.

### Generation of fluorescent *Mtb* reporter strain

Plasmid pVV16 containing hsp60 promoter and a gene encoding fluorescent protein mCherry (pVV16-mCherry plasmid, a gift from David Russell Lab) was transformed to *Mtb* by a Micropulser Electroporator (Biorad, USA). To prepare the competent cells for electroporation, an East Asia Beijing *Mtb* clinical strain isolated from a TB patient was cultured in 10 mL 7H9T medium at 37°C, with shaking, for 15 – 18 days to log phase OD_600_ 0.5 – 1. 1 mL of 2 M glycine solution was added to the culture 24 hours before harvesting the cells. Bacteria were harvested at 3200 g for 10 min, washed and re-suspended in 500 µL of 10% glycerol. 0.2-0.5 µg of pVV16-mCherry plasmid was mixed with 200 µL competent cells, left for 10 min, and then transferred to a 0.2 cm electroporation capped cuvette. Electroporation was conducted at a pulse of 2.5 kV, 25 µF. The cell suspension was transferred to a pre-warmed 7H9T medium, incubated at 37°C for at least 48 hours, and then plated on a selective medium containing 30 µg/mL of kanamycin. The plate was incubated at 37°C for 3 weeks. *Mtb* carrying the pVV16-mCherry plasmid were selected from pink colonies growing in the selective media and cultured in 7H9T media containing kanamycin at 37°C with shaking until OD_600_ 0.8 – 1 was reached. Bacteria were harvested, re-suspended in 10% glycerol, aliquoted and stored at −20°C for later use.

### Isolation of PBMC and human hMDM

Peripheral blood monocyte cells (PBMCs) were separated from 20 mL heparinized whole blood by Lymphoprep (Axis-Shield) gradient centrifugation in accordance with the manufacturer’s protocol and previous studies(Thuong et al. 2016). PBMCs were plated in cell-culture treated 60 mm × 15 mm petri dishes (Corning) with 6 – 8 ×10^6^ cells per dish in serum-free media and non-adhered cells were washed off. On the following day, for each sample, some cells were seeded in 96-well plates (8 ×10^4^ cells per well) and incubated at 5% CO_2_, 37°C in complete media (RPMI 1640 (Sigma) supplemented with 10% heat-inactivated fetal bovine serum (FBS, Sigma), 2mM L-glutamine (Sigma), 100 units of penicillin (Sigma)) containing 10 ng/mL human macrophage colony-stimulating factor (m-CSF, R&D system). Cells left over were cryopreserved in FBS + 10% DMSO for further infection experiments. To derive monocytes, adhered cells were incubated for 5 – 7 days. The complete media was changed at day 4, and the assays were performed at day 7.

### Preparation of acidification and proteolytic beads

Beads coated with ligands (IgG, TDM or β-glucan) to measure acidification and proteolysis activities were prepared as previously described(Yates and Russell 2008; Tram et al. 2019). Briefly, 500 ul carboxylated, 3 μm silica particles (Kisker Biotech) were washed and incubated at room temperature in 25 mg/mL cyanamide (Sigma-Aldrich) with 15 min agitation. Beads were then incubated with 1.0 mg defatted bovine serum albumin (Sigma-Aldrich) and 0.1 mg human immunoglobulin G (IgG, Molecular Probes), or 0.25 mg ligands [TDM (Enzo Life Sciences) or β-glucan/whole glucan particles (Invivogen)], together with DQ green BSA (2 mg/mL, Molecular Probes) in case of proteolysis for 12 hours with agitation. Coated beads were washed; for proteolysis, beads were then re-suspended in 10 μL of the 5 mg/mL stock of the calibration Alexa Fluor 594 SE, and agitated for 1 hour. For acidification activity, beads were labelled with the pH-sensitive reporter carboxyfluorescein-SE (10 µL of the 5 mg/mL stock, Molecular Probes) and agitated for 2 hours. Beads were then washed and re-suspended in 1 mL PBS with 0.02% sodium azide and stored at 4°C. The concentration of beads (number of beads per ml) was counted by FACS (BD).

### Acidification and proteolysis assays

The assays were performed as described previously(Tram et al. 2019). Briefly, macrophages were incubated in 100 µL pre-warmed assay buffer (1 mM CaCl2, 2.7 mM KCl, 0.5 mM MgCl2, 5 mM dextrose, 10 mM HEPES, 10% FBS in PBS). Beads were added into each well at a concentration sufficient to achieve an average of 4 – 5 beads internalized per macrophage as previously reported(Podinovskaia et al. 2013). This ratio of beads allows measurement of the change in fluorescent intensity in macrophages that corresponds to a drop of pH from 7.5 to 5.5, which signifies phagosomal maturation of macrophages at resting stage (Podinovskaia et al. 2013; Tram et al. 2019). Acidification and proteolysis assays were read for 90 min and 210 min respectively at 37°C in a fluorescent microplate reader (SynergyH4, BioTek). Data were collected at intervals of 1 min 30 s. The relative pH within phagosomes was reflected by the relative fluorescent unit (RFU) which is the ratio between fluorescent intensities emitted at 520 nm when excited at a pH-sensitive wavelength of 490 nm and at a pH-insensitive wavelength of 450 nm. Hydrolyzed DQ-BSA substrate emits at 520 nm when excited at 490 nm. Proteolysis activity was determined by the ratio between fluorescent intensities of substrate and calibration fluor. Activity index was used as a readout for macrophage acidification or proteolysis activity as reported(Tram et al. 2019) to access the variation in macrophage activities among individuals in response to different ligands or in different TB groups. The acidification activity index of macrophages at 30, 60, and 90 min was calculated by the ratio of the RFU at 10 min over that at 30, 60, or 90 min, respectively. Likewise, the proteolysis activity index at 60, 120, 180 and 210 min was calculated by dividing the RFU at these time-points by that at 10 min.

### Macrophage infection

*Mtb* reporter strain was cultured from −20°C stock in 7H9T containing 30 µg/mL kanamycin at 37°C with shaking for about 15 – 20 days until OD_600_ reached 1 – 2. The bacteria was then sub-cultured. When OD_600_ reached 0.5 – 1 the culture was used for macrophage infection.

Frozen monocytes from LTB, PTB and TBM were rapidly thawed at 37°C on the same day. Cells were then seeded in black flat bottomed 96-well plates at 8×10^4^ cells per well and differentiated to hMDM as described above. At day 7 after thawing, macrophages were infected with the fluorescent *Mtb* reporter at a multiplicity of infection (MOI) of 1, and incubated at 37°C, 5 % CO_2_. Four hours after infection, macrophages were washed with a pre-warmed medium without antibiotics. The intracellular growth of the *Mtb* reporter strain was assessed over 5 days using a fluorescent microplate reader. This measured the intensity of mCherry protein fluorescence at 620 nm after excitation at 575 nm. The viability of macrophages was observed under light microscopy.

### Statistical analysis

The donor-to-donor variation in phagosomal activities of macrophages from groups of healthy subjects or active TB patients was expressed by the percentage of coefficient of variation (% CV) calculated by the standard deviation (SD) divided by the mean. The difference in the variation of human macrophage activities across different ligands and TB groups were tested using the R package cvequality (Marwick, B. and Krishnamoorthy 2019). Comparisons between two groups for continuous variables or binary variables were performed using Mann–Whitney U-test or chi-squared test, respectively. P-values ≤0.05 were considered statistically significant. Graphs were generated by GraphPad Prism v7.03 or R program v3.3.1 (R Core Team, 2016).

## Results

### Characteristics of participants

The study participants comprised 53 healthy volunteers, of whom 35 had LTB infection, and 147 patients with active TB (ATB), consisting of 70 PTB and 77 TBM cases. The demographics of these groups are given in Table 1. The data for all characteristics were missing for 15 healthy volunteers, comprising 11 LTB and 4 TB uninfected individuals. The majority of ATB patients were male (71.4%), with a median age of 43 (IQR 32–50) while the group of healthy donors were 50% male with a median age of 38 (IQR 33.5–42.0). ATB patients had significantly higher absolute numbers of peripheral blood white blood cells, neutrophils and monocytes than healthy subjects (all P<0.0001). The number of lymphocytes of ATB patients was lower than that of healthy subjects (P=0.0009). Within the ATB group, PTB cases had significantly higher numbers of blood lymphocytes and monocytes than TBM patients (P≤0.0001, P=0.02 respectively).

**Table 1.**
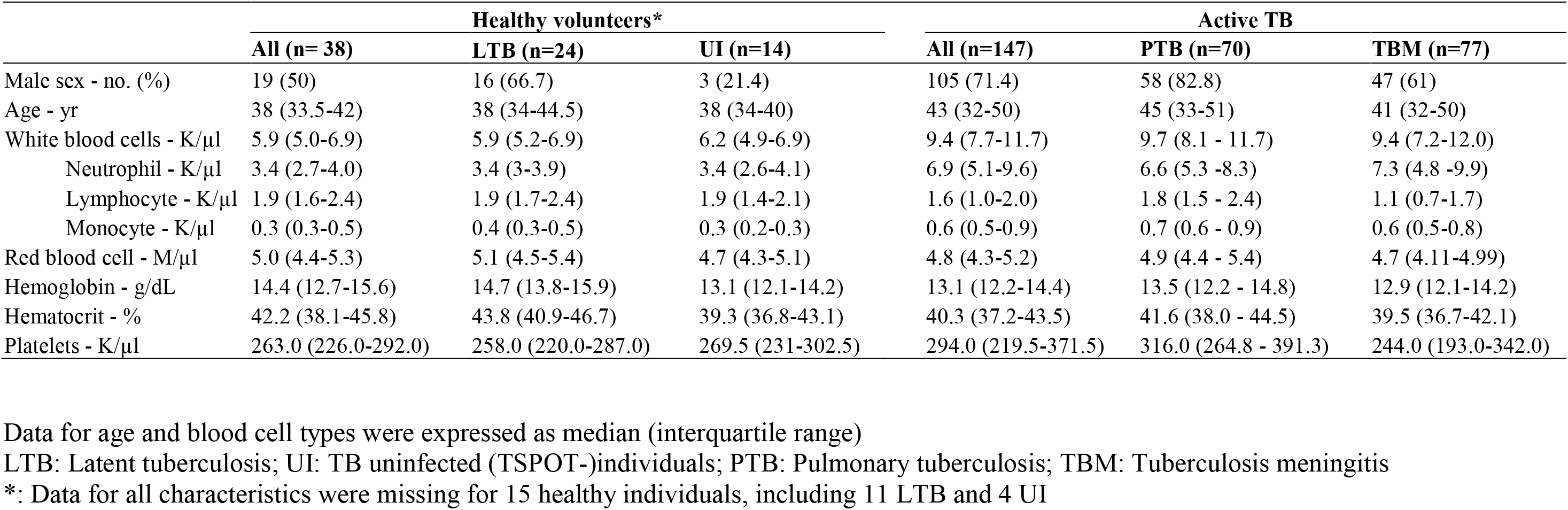
Baseline characteristics of study population

|  | Healthy volunteers* |  |  | Active TB |  |  |
| --- | --- | --- | --- | --- | --- | --- |
|  | All (n= 38) | LTB (n=24) | UI (n=14) | All (n=147) | PTB (n=70) | TBM (n=77) |
| Male sex - no. (%) | 19 (50) | 16 (66.7) | 3 (21.4) | 105 (71.4) | 58 (82.8) | 47 (61) |
| Age - yr | 38 (33.5-42) | 38 (34-44.5) | 38 (34-40) | 43 (32-50) | 45 (33-51) | 41 (32-50) |
| White blood cells - K/μl | 5.9 (5.0-6.9) | 5.9 (5.2-6.9) | 6.2 (4.9-6.9) | 9.4 (7.7-11.7) | 9.7 (8.1 - 11.7) | 9.4 (7.2-12.0) |
| Neutrophil - K/μl | 3.4 (2.7-4.0) | 3.4 (3-3.9) | 3.4 (2.6-4.1) | 6.9 (5.1-9.6) | 6.6 (5.3 -8.3) | 7.3 (4.8 -9.9) |
| Lymphocyte - K/μl | 1.9 (1.6-2.4) | 1.9 (1.7-2.4) | 1.9 (1.4-2.1) | 1.6 (1.0-2.0) | 1.8 (1.5 - 2.4) | 1.1 (0.7-1.7) |
| Monocyte - K/μl | 0.3 (0.3-0.5) | 0.4 (0.3-0.5) | 0.3 (0.2-0.3) | 0.6 (0.5-0.9) | 0.7 (0.6 - 0.9) | 0.6 (0.5-0.8) |
| Red blood cell - M/μl | 5.0 (4.4-5.3) | 5.1 (4.5-5.4) | 4.7 (4.3-5.1) | 4.8 (4.3-5.2) | 4.9 (4.4 - 5.4) | 4.7 (4.11-4.99) |
| Hemoglobin - g/dL | 14.4 (12.7-15.6) | 14.7 (13.8-15.9) | 13.1 (12.1-14.2) | 13.1 (12.2-14.4) | 13.5 (12.2 - 14.8) | 12.9 (12.1-14.2) |
| Hematocrit - % | 42.2 (38.1-45.8) | 43.8 (40.9-46.7) | 39.3 (36.8-43.1) | 40.3 (37.2-43.5) | 41.6 (38.0 - 44.5) | 39.5 (36.7-42.1) |
| Platelets - K/μl | 263.0 (226.0-292.0) | 258.0 (220.0-287.0) | 269.5 (231-302.5) | 294.0 (219.5-371.5) | 316.0 (264.8 - 391.3) | 244.0 (193.0-342.0) |

Data for age and blood cell types were expressed as median (interquartile range)
LTB: Latent tuberculosis; UI: TB uninfected (TSPOT-)individuals; PTB: Pulmonary tuberculosis; TBM: Tuberculosis meningitis
\*: Data for all characteristics were missing for 15 healthy individuals, including 11 LTB and 4 UI

### Variation of macrophage antimicrobial activities and their responses to immunogenic and non-immunogenic ligands

To understand the diversity of human macrophage activities among individuals and in response to different ligands, we examined the acidification and proteolysis of hMDM from 53 healthy subjects (35 with latent *Mtb* infection) using the reporter beads coated with the non-immunogenic IgG control antibody and with immunogenic ligands comprising TDM extracted from *Mtb* and β-glucan from *Saccharomyces cerevisiae*.

Macrophages treated with IgG, TDM or β-glucan displayed similar acidification kinetics: the phagosomal pH reduced significantly after 10-15 min of bead treatment and reached a plateau after 75-90 min (Figure 2A). Our previous study showed that for macrophages treated with IgG beads, such a change in acidification within 90 min corresponded to a decrease of phagosomal pH from 6.9 to 5.5(Tram et al. 2019). For all ligands, activity increased from the early time point of 30 min to 90 min when the phagosomes were completely acidified (Table 2). The activity varied among individuals, with ranges shown in Table 2. No significant difference was seen in the extent of variation of macrophage activity from these 53 subjects across IgG, TDM and β-glucan beads (Table 2) or from subgroups of LTB and uninfected individuals (Table S1 and S2). We then compared the macrophage acidification kinetics for the three ligands (Figure 2C). After 30, 60 and 90 min, IgG coated beads induced more acidification compared to other ligand coated beads (IgG vs. TDM beads: P=10^−5^, 3×10^−5^ and 0.01 at 30, 60 and 90 min respectively; IgG vs. β-glucan: P=10^−4^, 3×10^−4^ and 0.04 at 30, 60 and 90 min), but there was no statistical difference in activity between the beads coated with the two immunogenic ligands, TDM and β-glucan. The higher acidification activity of macrophages in response to IgG was also observed in both subgroups of LTB and uninfected persons categorized by T.SPOT (Figure S1).

**Table 2.**
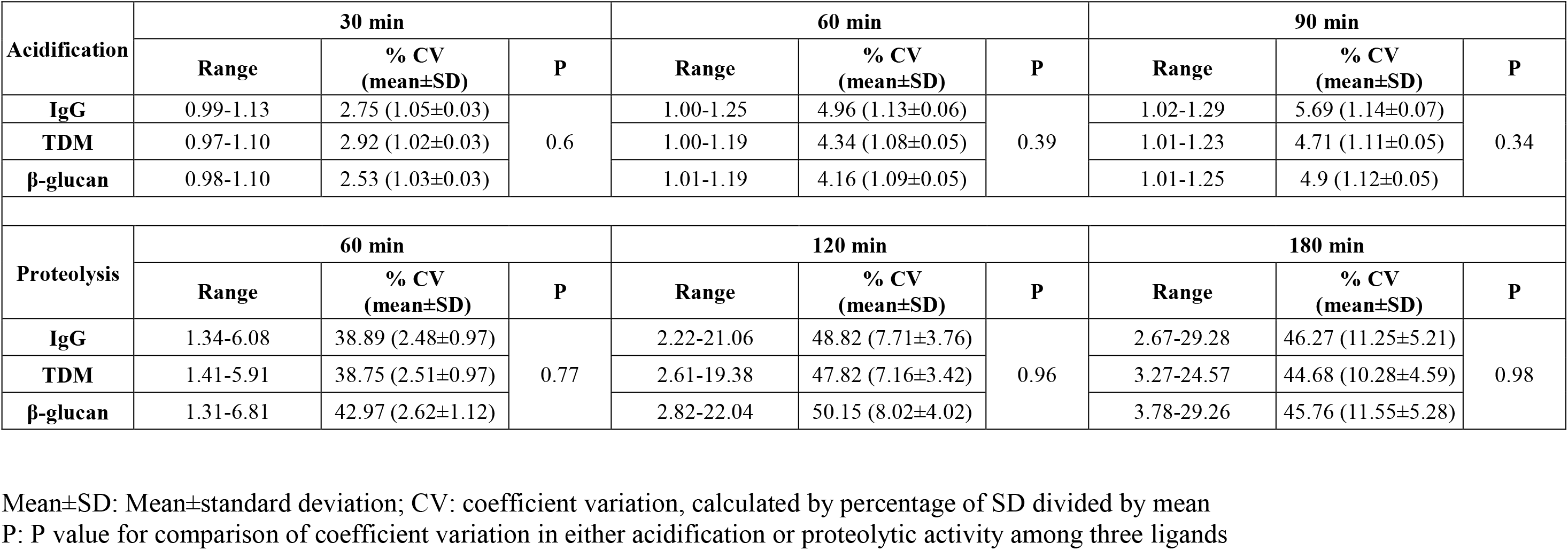
Variation in acidification and proteolytic activities in hMDM from healthy donors (n = 53) in response to IgG, TDM and β-glucan beads

| Acidification | 30 min |  |  | 60 min |  |  | 90 min |  |  |
| --- | --- | --- | --- | --- | --- | --- | --- | --- | --- |
|  | Range | % CV<br>(mean±SD) | P | Range | % CV<br>(mean±SD) | P | Range | % CV<br>(mean±SD) | P |
| IgG | 0.99-1.13 | 2.75 (1.05±0.03) | 0.6 | 1.00-1.25 | 4.96 (1.13±0.06) | 0.39 | 1.02-1.29 | 5.69 (1.14±0.07) | 0.34 |
| TDM | 0.97-1.10 | 2.92 (1.02±0.03) |  | 1.00-1.19 | 4.34 (1.08±0.05) |  | 1.01-1.23 | 4.71 (1.11±0.05) |  |
| β-glucan | 0.98-1.10 | 2.53 (1.03±0.03) |  | 1.01-1.19 | 4.16 (1.09±0.05) |  | 1.01-1.25 | 4.9 (1.12±0.05) |  |
| Proteolysis | 60 min |  |  | 120 min |  |  | 180 min |  |  |
|  | Range | % CV<br>(mean±SD) | P | Range | % CV<br>(mean±SD) | P | Range | % CV<br>(mean±SD) | P |
| IgG | 1.34-6.08 | 38.89 (2.48±0.97) | 0.77 | 2.22-21.06 | 48.82 (7.71±3.76) | 0.96 | 2.67-29.28 | 46.27 (11.25±5.21) | 0.98 |
| TDM | 1.41-5.91 | 38.75 (2.51±0.97) |  | 2.61-19.38 | 47.82 (7.16±3.42) |  | 3.27-24.57 | 44.68 (10.28±4.59) |  |
| β-glucan | 1.31-6.81 | 42.97 (2.62±1.12) |  | 2.82-22.04 | 50.15 (8.02±4.02) |  | 3.78-29.26 | 45.76 (11.55±5.28) |  |

Mean ± SD: Mean ± standard deviation; CV: coefficient variation, calculated by percentage of SD divided by mean P: P value for comparison of coefficient variation in either acidification or proteolytic activity among three ligands

Proteolytic activity, measured over 210 min, increased with time in a similar way for macrophages treated with IgG, TDM or β-glucan ligand-coated beads (Figure 2B). We used the normalized activity index at time points of 60, 120 and 180 min to examine the inter-donor and inter-ligand variation in activity. The macrophages from the 53 healthy individuals showed wide ranges of proteolytic activity in response to different ligand beads (Table 2). For example, after 180 min, the ranges of activity were 2.7-29.3, 3.3-24.6, 3.8-29.3 for IgG, TDM or β-glucan beads respectively. This variation between individuals was further indicated by large CVs of 45% when highest activity was reached, which was much greater than that of acidification (CVs of 5%). As for acidification, the variation in proteolytic activity from donor to donor was similar for all ligands (Table 2, S1 and S2). Comparison of the median values of the activity index for IgG, TDM and β-glucan beads showed no significant difference between non-immunogenic and immunogenic ligand coated beads at any time point (Figure 2D).

In summary, our results showed variations in macrophage antimicrobial activities among healthy individuals, particularly for proteolysis. The extent of these variations was highly consistent across stimulation with different ligands. Meanwhile human macrophage acidification activity was higher in response to beads coated with non-immunogenic IgG than with pathogenic TDM or β-glucan.

### Variation of macrophage antimicrobial activities in LTB versus ATB

We next compared the macrophage antimicrobial activities in participants with LTB and ATB (Figure 1). Beads coated with TDM from *Mtb* were used to measure the activities of hMDM from 101 ATB and 35 LTB participants. Macrophages from both groups showed similar patterns of phagosomal acidification and proteolysis (Figure S2). The variation in acidification was not significantly different for these two groups (Table 3). Meanwhile for proteolytic activity the variation was greater in LTB than in ATB and the difference became significantly with time (at 120 min, P = 0.08; at 180 min CV = 40% in LTB, 29% in ATB, P = 0.03). The macrophages from TB uninfected individuals showed more variation than those from LTB or ATB (Table S3 and S4).

**Table 3.** Variation in acidification and proteolytic activities in hMDM from LTB (n=35) and ATB (n=101) in response to TDM beads

| Acidification | 30 min |  |  | 60 min |  |  | 90 min |  |  |
| --- | --- | --- | --- | --- | --- | --- | --- | --- | --- |
|  | Range | % CV<br>(mean±SD) | P | Range | % CV<br>(mean±SD) | P | Range | % CV<br>(mean±SD) | P |
| LTB | 0.97-1.10 | 3.12 (1.02±0.03) | 0.32 | 1.00-1.19 | 3.97 (1.09±0.04) | 0.41 | 1.02-1.23 | 4.53 (1.12±0.05) | 0.39 |
| ATB | 0.98-1.10 | 2.75 (1.03±0.03) |  | 1.00-1.23 | 4.52 (1.1±0.05) |  | 1.00-1.32 | 5.14 (1.14±0.06) |  |
| Proteolysis | 60 min |  |  | 120 min |  |  | 180 min |  |  |
|  | Range | % CV<br>(mean±SD) | P | Range | % CV<br>(mean±SD) | P | Range | % CV<br>(mean±SD) | P |
| LTB | 1.45-5.91 | 36.53 (2.71±0.99) | 0.24 | 2.80-19.38 | 42.87 (8±3.43) | 0.08 | 3.53-24.57 | 39.98 (11.42±4.57) | 0.03 |
| ATB | 1.00-4.85 | 30.61 (2.91±0.89) |  | 1.74-15.3 | 32.95 (8.56±2.82) |  | 2.31-21.27 | 29.1 (12.12±3.53) |  |

LTB: Latent tuberculosis; ATB: Active tuberculosis
Range: min-max; Mean±SD: Mean±standard deviation; CV: coefficient variation, calculated by percentage of SD divided by mean
P: P value for comparison of coefficient variation in either acidification or proteolytic activity among two groups

**Figure 1.**
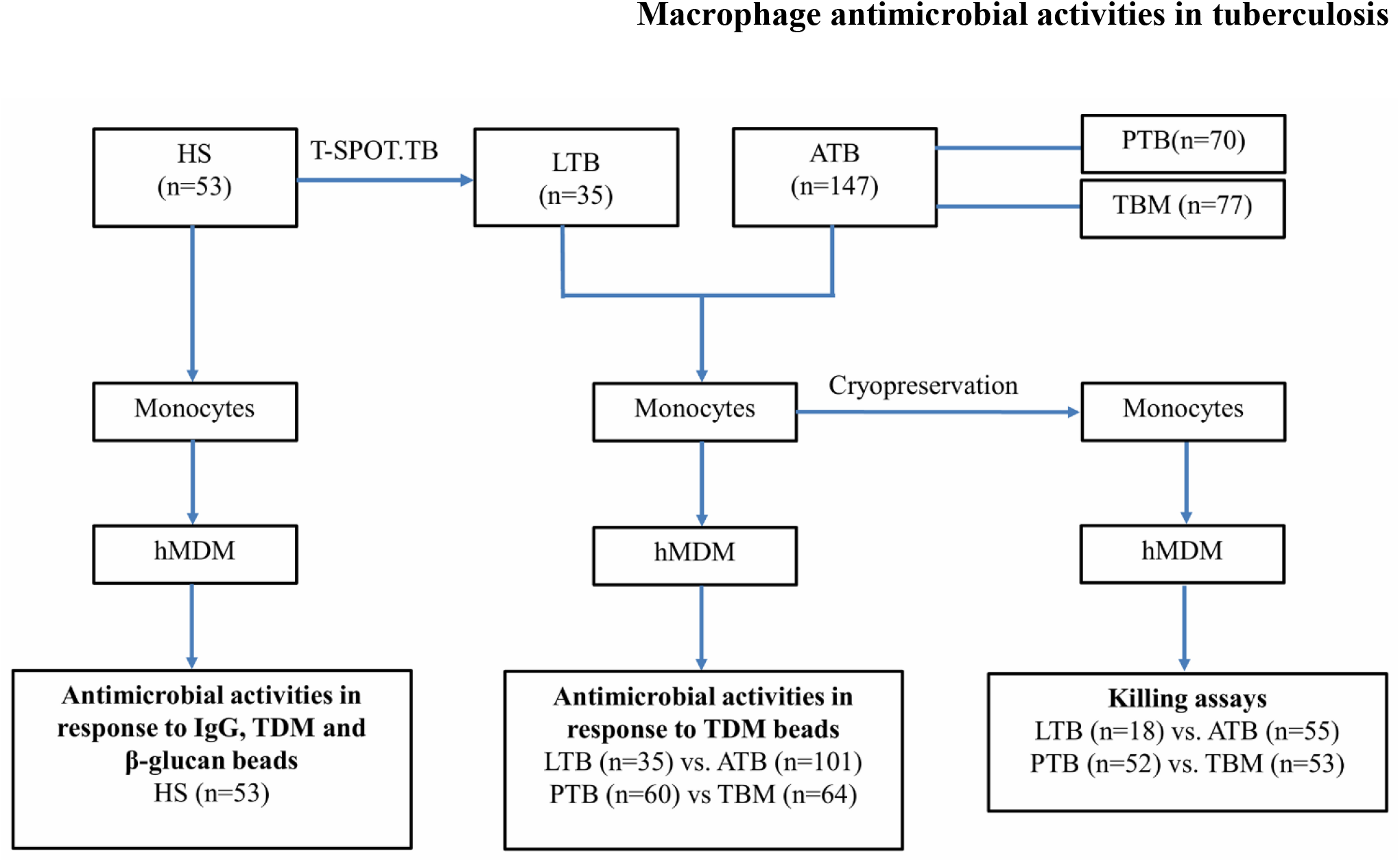
Study design. HS: Healthy volunteers; LTB: Latent tuberculosis; ATB: Active tuberculosis; PTB: Pulmonary tuberculosis: TBM: Tuberculosis meningitis

**Figure 2.**
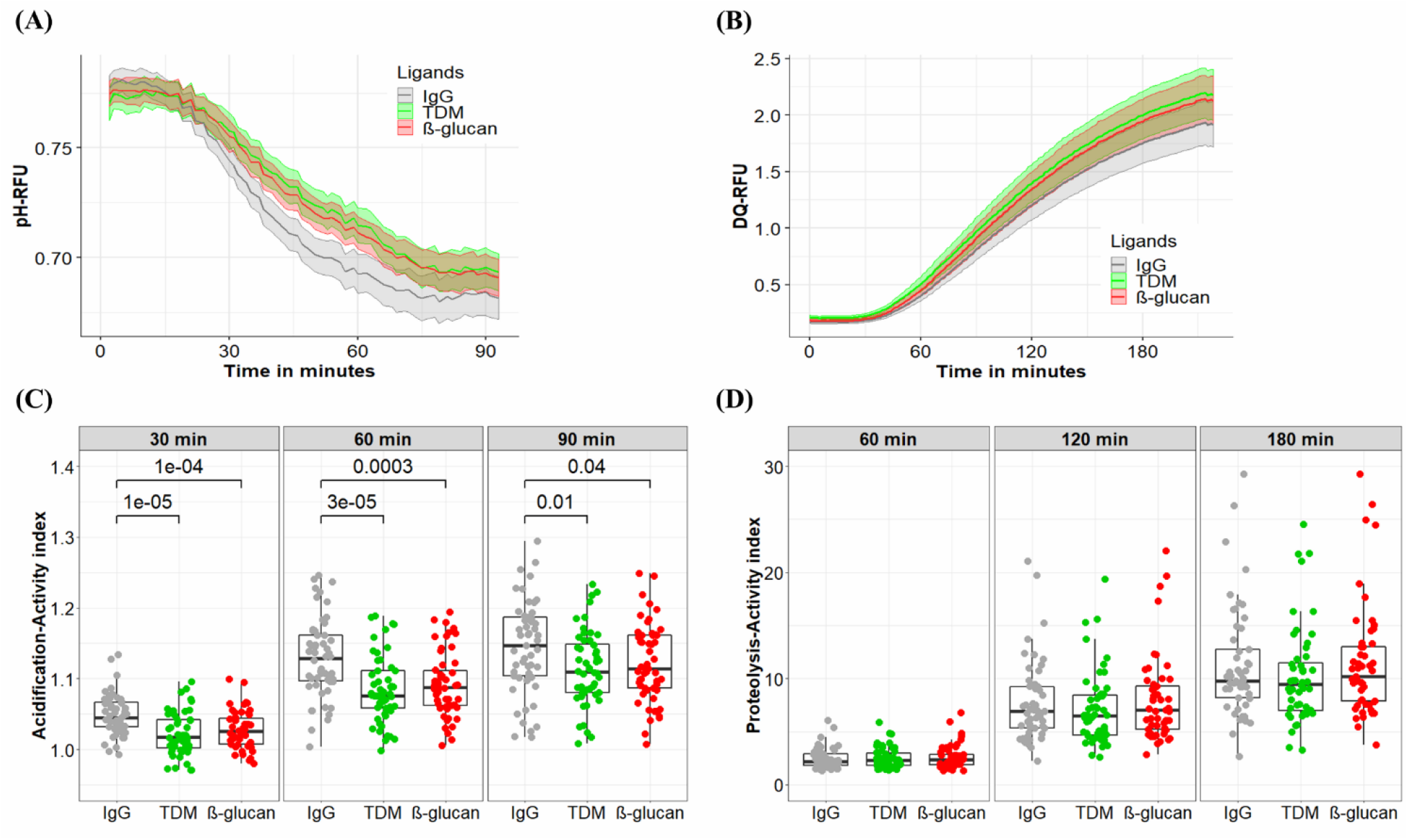
Antimicrobial activities of macrophages treated with different ligand beads. hMDM from healthy volunteer subjects (n = 53) at day 7 were treated with beads coated with either IgG, TDM or β-glucan to measure acidification activity for 90 min (A, C) or proteolytic activity for 180 min (B, D). (A) The kinetics of acidification of hMDM. (B) The kinetics of proteolysis of hMDM.The acidification activity index of hMDM at 30, 60, and 90 min. (D) The proteolysis activity index of hMDM at 60, 120, and 180 min. In (A, B) the line and the shaded area represent the mean activities and 95% confidence interval. In (C, D) box plots represent the interquartile range (IQR) and median. Vertical lines above and below each box extend to the most extreme data point that is within 1.5x IQR. Each dot represents the activity index of hMDM from each subject. P values were determined by Mann-Whitney U test.

We also compared median values of acidification and proteolytic activities in LTB versus ATB and found no difference at the time points measured (Figure 3A). These activities, particularly proteolysis, were significantly higher in either LTB or ATB in comparison to 14 healthy subjects who were negative with T.SPOT (Figure S3). Ability to control bacterial growth was assessed for macrophages from 18 LTB and 55 ATB patients both by measuring mCherry intensity, indicative of *Mtb* survival (Figure S4) and by observing the viability of infected macrophages. For both groups, bacterial survival gradually increased with time (Figure 3B) while macrophage viability gradually decreased (Figure 3C). Comparisons between LTB and ATB groups showed no difference in either mCherry intensity or cell lysis during 5 days of infection.

**Figure 3.**
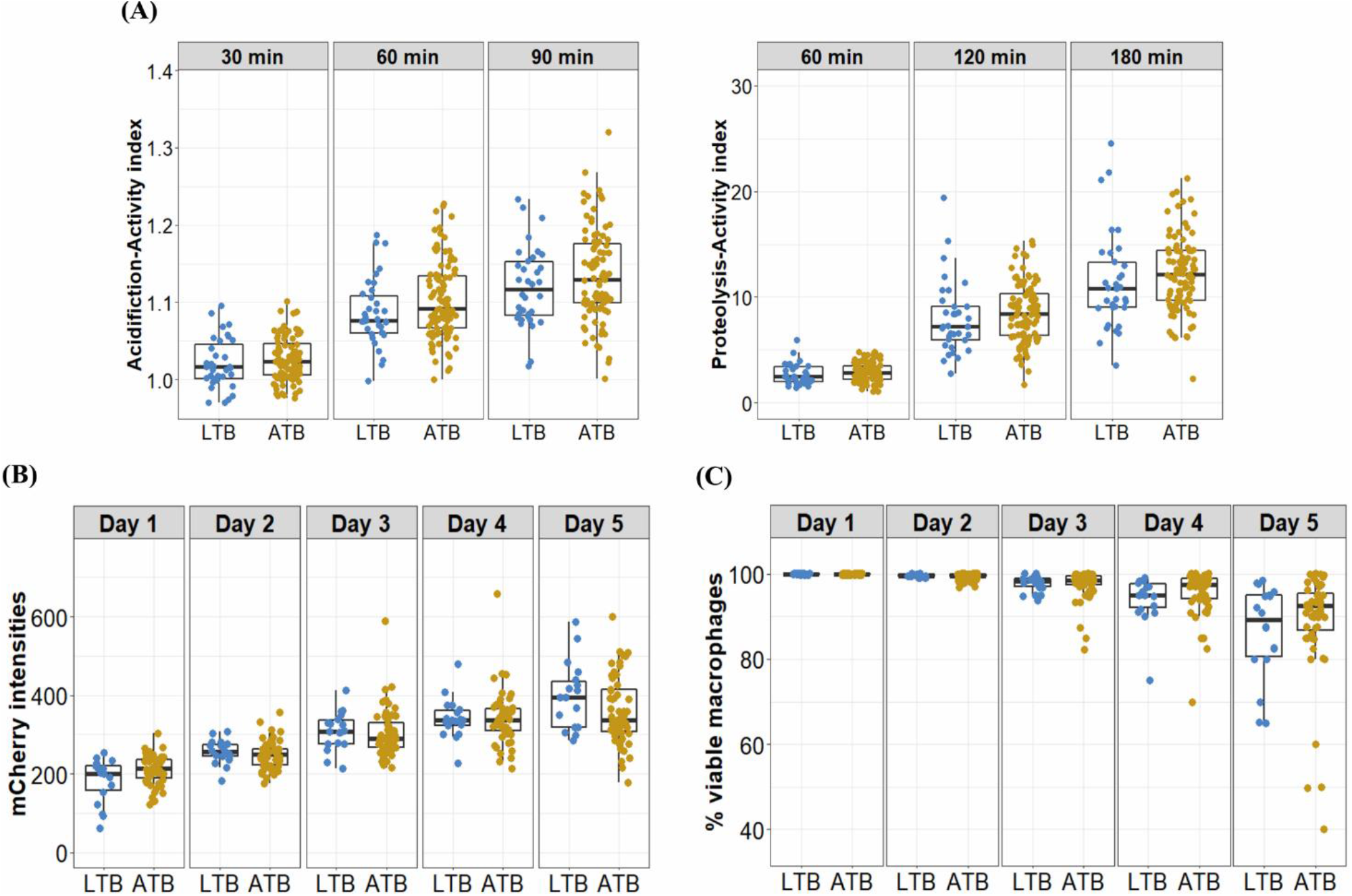
Antimicrobial activities and bacterial control of macrophages from LTB and ATB participants. (A) hMDM from LTB (n = 35) and ATB (n = 101) participants at day 7 were treated with beads coated with TDM to measure acidification activity after 30, 60 and 90 min, or proteolytic activity after 60, 120 and 180 min. (B, C) Macrophages derived from cryopreserved monocytes from LTB (n = 18) or ATB (n = 55) at day 7 were infected with a *Mtb* reporter strain at MOI 1. The bacterial intracellular growth during 5 days of infection was assessed by the mCherry intensity readout (B) and the viability of infected macrophages (C). Box plots represent the interquartile range (IQR) and median. Vertical lines above and below each box extend to the most extreme data point that is within 1.5x IQR. Each dot represents the activity index of hMDM from each subject.

### Relationship between macrophage antimicrobial activities and disseminated tuberculosis

Acidification and proteolytic activities and ability to control bacteria of macrophages from patients with TBM (n= 64) were compared with those from PTB patients (n=60) (Figure 1). The variation in macrophage acidification and proteolytic activities in response to the TDM-coated beads looked similar for TBM patients as for PTB patients (Table 4, Figure 4A). At all time points in acidification and proteolysis, there was no significant difference in the activity levels between the two groups of patients. *Mtb* survival and viability of infected macrophages were also assessed for macrophages from 52 PTB and 53 TBM patients. For both PTB and TBM we observed an increased mCherry signal in infected macrophages during 5 days of infection (Figure 4B). All infected macrophages remained adherent and intact until day 3 when some of the cells became lysed (Figure 4C). The hMDM from two groups of patients showed no difference in either mCherry intensity or cell lysis during the 5 days.

**Table 4.** Variation in acidification and proteolysis activities in hMDM from PTB (n=60) and TBM (n=64) in response to TDM beads

| Acidification | 30 min |  |  | 60 min |  |  | 90 min |  |  |
| --- | --- | --- | --- | --- | --- | --- | --- | --- | --- |
|  | Range | % CV<br>(mean±SD) | P | Range | % CV<br>(mean±SD) | P | Range | % CV<br>(mean±SD) | P |
| PTB | 0.98-1.09 | 2.69 (1.03±0.03) | 0.78 | 1.02-1.23 | 4.59 (1.11±0.05) | 0.66 | 1.02-1.32 | 4.83 (1.14±0.06) | 0.77 |
| TBM | 0.98-1.1 | 2.77 (1.03±0.03) |  | 1-1.22 | 4.3 (1.1±0.05) |  | 1-1.27 | 5.01 (1.13±0.06) |  |
| Proteolysis | 60 min |  |  | 120 min |  |  | 180 min |  |  |
|  | Range | % CV<br>(mean±SD) | P | Range | % CV<br>(mean±SD) | P | Range | % CV<br>(mean±SD) | P |
| PTB | 1.31-5.4 | 29.3 (2.97±0.87) | 0.28 | 3.59-14.96 | 32.08 (8.47±2.72) | 0.35 | 6.14-19.74 | 28.51 (11.81±3.37) | 0.35 |
| TBM | 1.09-4.85 | 34.2 (2.79±0.95) |  | 1.74-15.30 | 37.13 (8.21±3.05) |  | 2.31-21.27 | 32.85 (11.78±3.87) |  |

PTB: Pulmonary tuberculosis; TBM: Tuberculosis meningitis
Range: min-max; Mean±SD: Mean±standard deviation; CV: coefficient variation, calculated by percentage of SD divided by mean
P: P value for comparison of coefficient variation in either acidification or proteolytic activity among two groups

**Figure 4.**
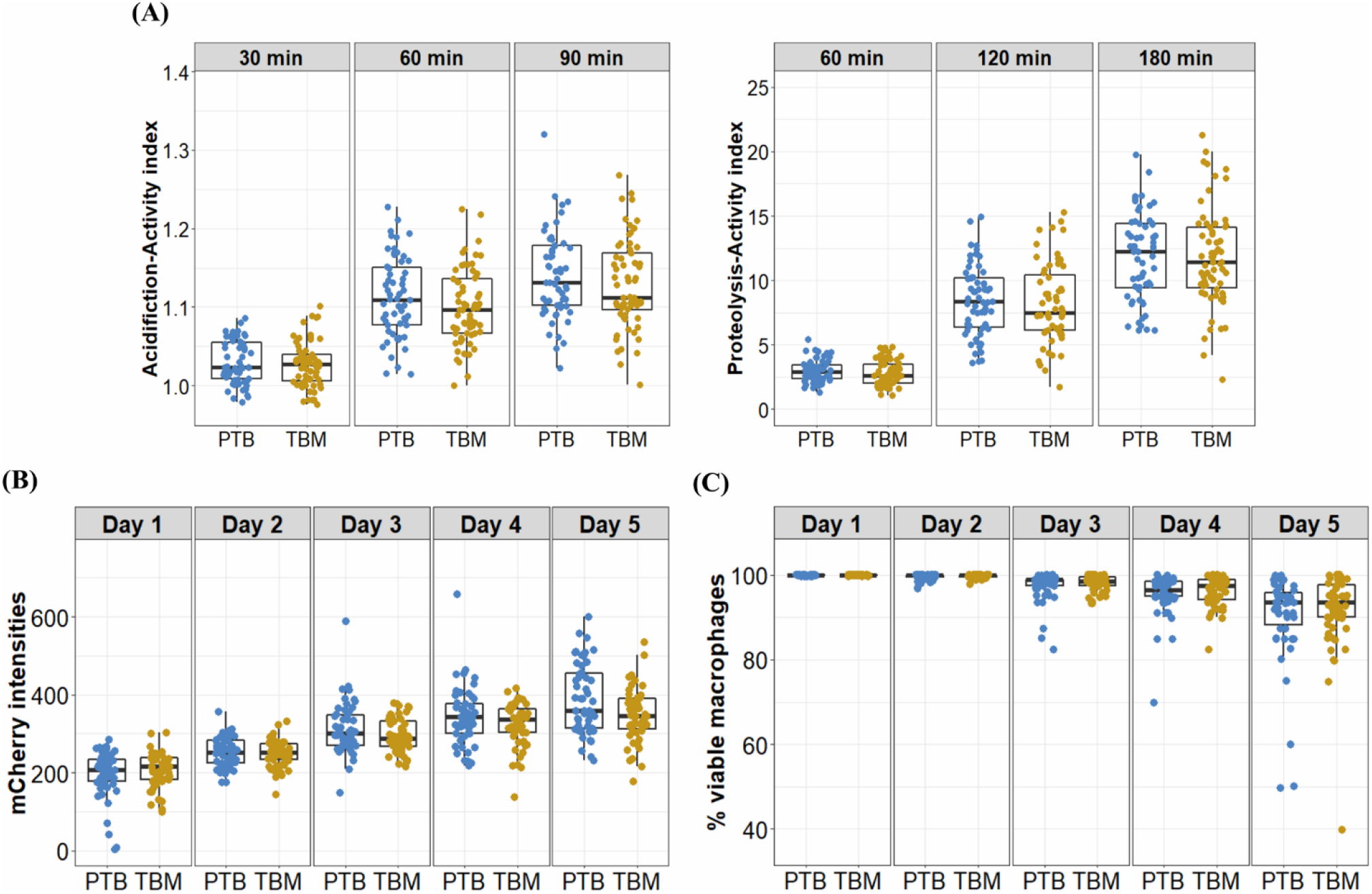
Antimicrobial activities and bacterial control of macrophages from PTB and TBM participants. (A) hMDM from patients with either PTB (n = 60) or TBM (n = 64) at day 7 were treated with TDM-coated beads, to measure acidification activity after 30, 60 and 90 min, or proteolytic activity after 60, 120 and 180 min. (B, C) Macrophages derived from cryopreserved monocytes from patients with PTB (n = 52) or TBM (n = 53) at day 7 were infected with a *Mtb* reporter strain at MOI 1. The bacterial intracellular growth during 5 days of infection was assessed by the mCherry intensity readout (B) and the viability of infected macrophages (C). Box plots represent the interquartile range (IQR) and median. Vertical lines above and below each box extend to the most extreme data point that is within 1.5x IQR. Each dot represents the activity index of hMDM from each subject.

## Discussion

Macrophages are essential to innate immunity, controlling *Mtb* infection by activating the antimicrobial program and particularly performing killing reactions including acidification and proteolysis(Welin et al. 2011; Pires et al. 2016). In this study, we investigated antimicrobial activities of macrophages from people who were healthy, had latent TB or active TB, either PTB or TBM, to extend understanding of the impact of macrophage activities on TB disease protection and progression.

We explored person-to-person variations of acidification and proteolysis activities of macrophages stimulated with immunogenic or non-immunogenic ligands. These variations were highly preserved among different stimuli or among cells originating from different hosts, ranging from healthy to having different forms of TB. We found acidification activity of healthy donors’ macrophages was greater in response to non-immunogenic IgG than to pathogenic ligands TDM or β-glucan. We did not find any differences in observed activities between LTB and ATB, or PTB and TBM.

The adapted bead-based assays allowed us to investigate ligand-specific activities of macrophages. Acidification responses to ligand stimulation occurred rapidly and reduced pH in the phagosomal environment from neutral to below 5.5, which then facilitates later hydrolytic activity such as proteolysis. In the healthy population, we found the variation of proteolysis (CV 45%) to be much greater than that of acidification (CV 5%). However, for both activities, the variations remained unchanged when macrophages were activated with either non-immunologic or pathogenic ligands, suggesting a stable diversity of macrophage activities among different individuals in response to various stimuli. Although inter-donor variations of these activities were preserved for different ligands, acidification activities were much stronger for macrophages stimulated with IgG than with either TDM or β-glucan. These findings, together with those of Podinovskaia *et al*,(Podinovskaia et al. 2013) highlight that macrophage intracellular activities at the level of individual phagosomes and individual persons are specific to the nature of stimuli. Macrophages treated with non-immunogenic IgG, recognized by Fc-gamma receptor 1, represent a resting stage and maintain homeostasis, hence they perform acidification and proteolytic activities strongly for regular removal of cell debris or apoptotic cells(Podinovskaia et al. 2013). TDM and β-glucan are pathogenic ligands recognized by macrophage receptors MARCO, Mincle and Dectin-1(Ishikawa et al. 2009; Bowdish et al. 2009; Brown and Gordo 2001), activating the innate immune response that could lead to a modulation of macrophage acidification for efficient antigen processing(Russell et al. 2009) or for better pathogen intracellular survival. In line with the latter possibility, previous studies in BMDM show that TDM inhibits phagosome-lysosome fusions and reduces acidification to enhance intracellular survival of mycobacterium during infection(Indrigo, Hunter, and Actor 2003; Fineran et al. 2017).

TDM is one of the most abundant glycolipids on the *Mtb* surface and induces the innate macrophage response to the infection(Indrigo, Actor, and Hunter 2002). Beads coated with TDM exhibit a similar trafficking pattern to the live virulent Erdman *Mtb* in mice macrophages(Indrigo, Hunter, and Actor 2003). Work in human macrophages shows delayed acidification and proteolysis activities in macrophages infected with *Mtb* compared to uninfected macrophages when using reporter beads coated with IgG (Podinovskaia et al. 2013). In our study, acidification and proteolysis activities of macrophages treated with TDM coated beads were lower than for IgG coated beads. Although other *Mtb* ligands could also influence results by interacting with different host pattern recognition receptors, all these results together suggest similar patterns in antimicrobial activities of macrophages in response to beads coated with TDM and live *Mtb*.

In both LTB and ATB groups, individuals showed heterogeneity in their macrophage antimicrobial activities. The variation of macrophage proteolysis in LTB was slightly greater than the variation in ATB, indicating wider capacity of this activity and thus its potential involvement in protecting healthy subjects from developing active TB. Although the mean values of measured activities were not significantly different in LTB and ATB, such diversity in proteolysis and acidification among individuals allows for further study for underlying factors such as genetic variants in the phagocytic genes which initiate the phagocytosis and phagosomal activities of macrophages(Thuong et al. 2016).

Antimicrobial activities of macrophages are important for controlling bacterial replication and could be involved in bacterial dissemination from one locality to other organs. In this study, macrophages from PTB and TBM showed similarity in their levels of antimicrobial activity as well as in their ability to control *Mtb*, suggesting these activities are not associated with *Mtb* spreading in patients.

The *ex vivo* human MDM model used in this study allowed us to examine primary macrophages from many subjects, from those without infection to those with different TB phenotypes, but also has some limitations. Firstly, our model could not reflect the heterogeneity of lung macrophage populations during the course of *Mtb* infection, which may influence the outcome of infections. Secondly, using a real-time fluorescence plate reader, we could only measure phagosome acidification and bulk protease activity, not other hydrolysis and oxidative activity(Brown and Gordo 2001). The antimicrobial activities of macrophages are also known to be modulated by antibodies or neutrophils (Lu et al. 2016; B. H. Tan et al. 2006; Lu et al. 2020), which cannot be captured in a macrophage model.

Our study is the first to investigate the variation of antimicrobial activities of human MDM from different individuals and their association with different TB clinical manifestations. Antimicrobial activities of macrophages were found to vary among individuals and show immunological dependence. However, our results suggest that these activities cannot be solely responsible for the control of bacterial replication or dissemination in TB.

## Data Availability

The authors confirm that the data supporting the findings of this study are available within the article and its supplementary materials.

## Acknowledgments

We thank the participants in this study, and the clinicians and nurses at HTD and DTU 4 and 8 who helped to perform this study. We would like to thank David Russell from Cornell University for provision of pVV16-mCherry plasmid, and to Vijay Srinivasan for his critical reading of this manuscript.

## Conflict of Interest

*The authors declare that the research was conducted in the absence of any commercial or financial relationships that could be construed as a potential conflict of interest*.

## Author Contributions

TTBT, NTTT, NHP and GT conceived and designed the experiments. TTBT, VTNH, DDAT, TDD, HNN, NTH did the experiments and collected the data. TTBT, GT and NTTT analyzed and interpreted the data. TTBT, VTNH, DDAT, TDD, HNH, NTH, NHP, GT and NTTT drafted, revised the manuscript and approved the final version.

## Funding

This work was supported by Wellcome Trust Fellowship in Public Health and Tropical Medicine to NTTT (097124/Z/11/Z and 206724/Z/17/Z) and Wellcome Trust Major Overseas Program Funding to GT (106680/B/14/Z).

## Notes

### Competing Interest Statement

The authors have declared no competing interest.

### Author Declarations

The study protocol and informed consent form were approved by the institutional review boards of the Hospital for Tropical Diseases in Vietnam and the Oxford Tropical Research Ethics Committee in the United Kingdom.

